# Modelling the cost-effectiveness of essential and advanced critical care for COVID-19 patients in Kenya

**DOI:** 10.1101/2021.08.16.21261894

**Authors:** Angela Kairu, Vincent Were, Lynda Isaaka, Ambrose Agweyu, Samuel Aketch, Edwine Barasa

## Abstract

**Background:** Case management of symptomatic COVID-19 patients is a key health system intervention. The Kenyan government embarked to fill capacity gaps in essential and advanced critical care needed for the management of severe and critical COVID-19. However, given scarce resources, gaps in both essential and advanced critical care persist. This study assessed the cost-effectiveness of investments in essential and advanced critical care to inform the prioritization of investment decisions.

**Methods:** We employed a decision tree model to assess the incremental cost-effectiveness of investment in essential care (EC) and investment in both essential and advanced critical care (EC+ACC) compared to current health care provision capacity (status quo) for COVID-19 patients in Kenya. We used a health system perspective, and an inpatient care episode time horizon. Cost data was obtained from primary empirical analysis while outcomes data was obtained from epidemiological model estimates. We used univariate and probabilistic sensitivity analysis (PSA) to assess the robustness of the results.

**Results:** The status quo option is more costly and less effective compared to investment in essential care and is thus dominated by the later. The incremental cost effectiveness ratio (ICER) of Investment in essential and advanced critical care (EC+ACC) was US $1,378.21 per DALY averted and hence not a cost-effective strategy when compared to Kenya’s cost-effectiveness threshold (USD 908).

**Conclusion:** When the criterion of cost-effectiveness is considered, and within the context of resource scarcity, Kenya will achieve better value for money if it prioritizes investments in essential care before investments in advanced critical care. This information on cost-effectiveness will however need to be considered as part of a multi-criteria decision-making framework that uses a range of criteria that reflect societal values of the Kenyan society.

**Key questions:** *What is already known?:*

- The COVID-19 pandemic is responsible for substantial health effects in low- and middle-income countries
- The case management of COVID-19 is one of the key control interventions deployed by country health systems.
- Similar to other low- and middle-income countries, Kenya had substantial gaps in both essential and advanced critical care at the beginning of the pandemic.

*What are the new findings?:*

- Provision of essential care and advanced critical care for COVID-19 at the current health system capacity (status quo) was costly and the least effective strategy.
- Investment in both essential care and advanced critical care for COVID-19 is not cost-effective in Kenya when compared to investment in essential care.

*What do the new findings imply?:*

- Prioritizing investments in filling capacity gaps in essential care before investing in filling capacity gaps in advanced critical care for COVID-19 is more cost-effective in Kenya
- These findings are intended to inform the sequencing of investments in case management rather than the selection of either strategy, within a context of substantial resource constraint, and capacity gaps in both essential and advanced critical care or COVID-19
- Kenya will need to consider these findings on cost-effectiveness within a multi-criteria decision-making framework that use a range of criteria that reflect societal values.

## INTRODUCTION

The new coronavirus disease 2019 (COVID-19) has spread to nearly all countries and territories globally, with devastating impacts (1). As at 20^th^ May 2021, 164.9 million cases of SARS-CoV-2 infections have been recorded resulting in 3.4 million deaths globally (2). In Kenya, 166,382 infections and 3,035 deaths from COVID-19 have been recorded as of 20^th^ May 2021 (2). Beyond direct health impacts, the COVID-19 pandemic is responsible for substantial indirect health effects that include the disruption of the delivery and access of routine health services. It is also responsible for negative socio-economic impacts that include a slow-down of the global economy, increase in unemployment, impoverishment, disruption of schooling and threatening of food security, among others (3).

The case management of COVID-19 is one of the key control interventions deployed by country health systems. COVID-19 is a highly contagious infectious disease transmitted by severe acute respiratory syndrome coronavirus 2 (SARS-CoV-2) primarily via exposure to respiratory droplets (1). Clinically, COVID-19 presents as either of four severities namely 1) asymptomatic, 2) Mild/Moderate, 3) Severe, and 4) Critical COVID-19 (4, 5). In Kenya, case management guidelines recommend asymptomatic and mild/moderate COVID-19 be managed at home (home based care), while patients with severe and critical COVID-19 are provided institutional care in hospitals (6, 7). Patients with severe COVID-19 are typically managed in general hospital wards, and receive essential care that may include supplemental oxygen support (1), whereas patients with critical COVID-19 are managed in intensive care units (ICUs) and provided with advanced critical care such as mechanical ventilation, management of complications like respiratory failure, acute respiratory distress syndrome (ARDS), thromboembolism, sepsis and septic shock, and multi-organ failure such as cardiac and acute kidney injury, provided in intensive care unit (ICU) (1, 8, 9).

Like other low- and middle-income countries (LMICs), Kenya had substantial gaps in both essential and advanced critical care at the beginning of the pandemic. For instance, it is estimated that only 58% of hospital beds had access to medical oxygen at the start of the pandemic (10). Further, only only 16% of healthcare facilities in Kenya were able to monitor oxygen saturation and therapy through pulse oximetry, and the mean availability of tracer items for emergency breathing interventions (pulse oximeters, micronebulizer, beclomethasone and salbutamol inhalers, oxygen with tubing, flowmeter, and humidifier, resuscitation bags, intubation devices with connecting tube, chest tubes with insertion sets, and CPAP equipment) was only 13% (10).

With regard to advanced critical care, Kenya had only 540 ICU beds for a population of nearly 50 million, with only 22% of the population living within 2 hours of a facility with an ICU (10).

Against this backdrop, the Kenyan government set out to invest in filling capacity gaps in both essential care and advanced critical care for COVID-19. However, because of resource constraints, gaps in both essential and advanced care persist one year into the pandemic (11). This has triggered discussions on the prioritization of investments when resources are scarce. If, as it is evident, the Kenyan government is unable to fill capacity gaps in both essential and advanced critical care, where should they start? In this paper, we use carry out an economic evaluation to inform this decision. Specifically, we compare the cost-effectiveness of investments in essential care and investments in advanced care in addition to essential care to the current health care provision capacity (status quo) for the management of symptomatic COVID-19 patients with severe and critical disease in Kenya.

## METHODS

### Study design

A decision tree analysis model in Tree Age Pro Healthcare 2020 was developed to evaluate the cost-effectiveness of investments in essential and advanced critical care for the management of COVID-19 patients in Kenya from a health systems perspective. The model followed a cohort of 2,288 individuals, representative of all individuals hospitalized for COVID-19 illness (until 30^th^ January 2021) through two treatment pathways, estimating costs and health gains. In both treatment pathways, the COVID-19 patients were diagnosed as either having severe or critical illness depending on the severity of symptoms as defined by the Kenya ministry of health COVID-19 case management guidelines (6). A diagnosis was followed by treatment of severe patients in a general ward and critical patients in the intensive care unit (ICU). A time horizon of a patient care episode chosen.

### Model structure

Three different treatment strategies are compared (Figure 1). Strategy one is defined as investment in filling essential care (EC) gaps broadly comprising of supplementary oxygen therapy (when needed), administration of empiric antimicrobials, monitoring of vital signs and laboratory tests. Strategy two is defined as investment in advanced critical care in addition to essential care (EC +ACC). Advanced critical care encompasses management of patients in an intensive care unit, advanced oxygen/ventilatory support, conservative fluid management, advanced organ monitoring and support, empiric antimicrobials, and management of any complications (6). Strategy three is the baseline defined as status quo comprising of provision of EC and ACC within the current health system capacity. Appendix 1 provides a detailed description of what is included in each intervention, which were defined based on expert consensus and the clinical guidelines implemented (1, 6).

**Figure 1:**
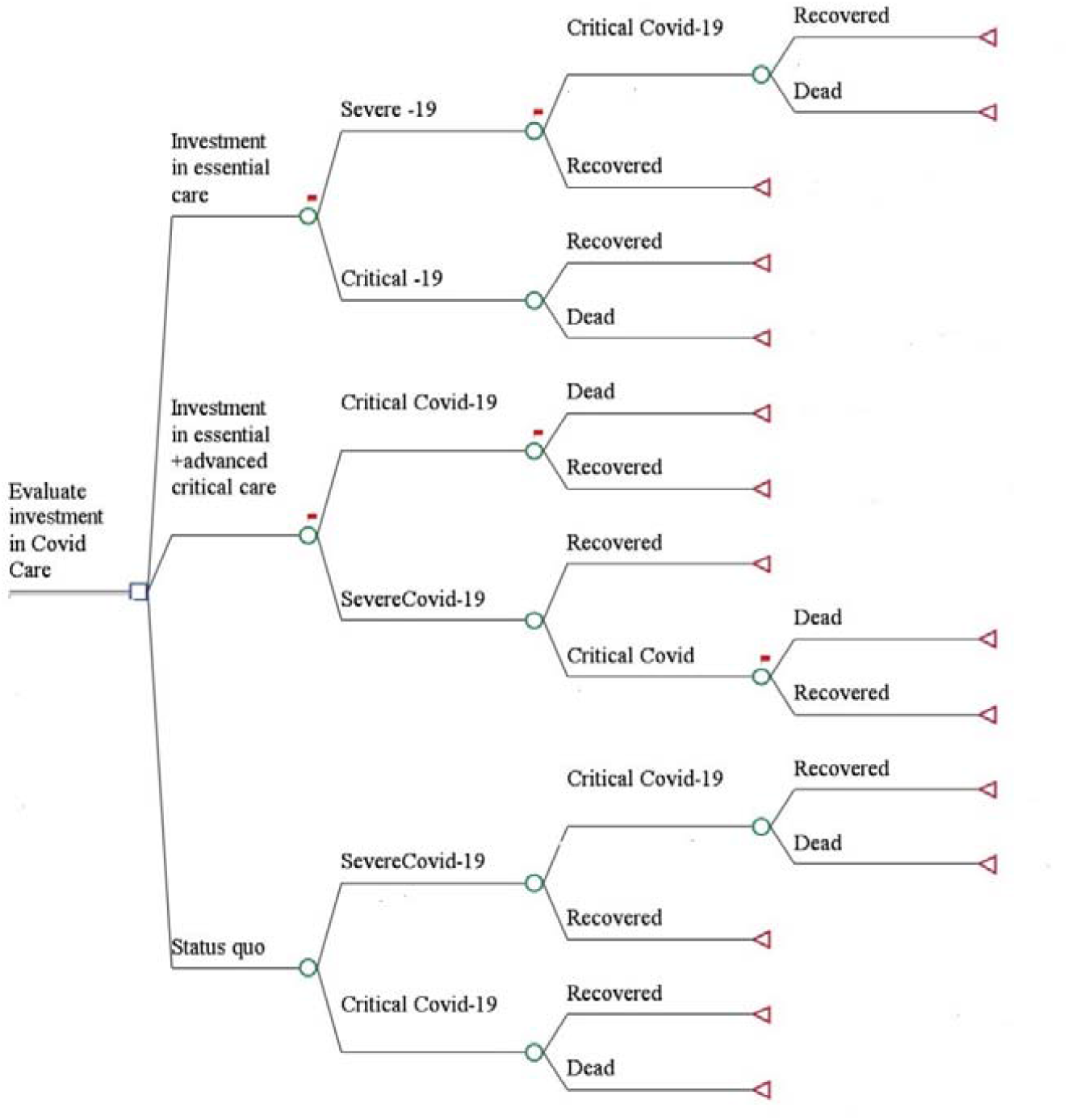
Schematic of decision tree model

The study population is hospitalized COVID-19 patients admitted between March 2020 to January 2021. The model assumes, in line with Kenya COVID-19 case management guidelines, that only patients with severe and critical disease are admitted in hospitals for inpatient care (6). Severe cases present with the following symptoms: fever or suspected respiratory infection, plus one of respiratory rate >30 breaths/min, severe respiratory distress, or SpO2 <93% (6). Critical cases are those who meet any of the following criteria: respiratory failure requiring mechanical ventilation, shock, and other organ failure requiring ICU care (6). All individuals completed the pathway when they were either recovered or dead. Data on the effectiveness of both comparators was obtained from current literature, and where unavailable assumptions were based on expert opinion. However, these data sources were limited by the regions/areas studied as the extent of disease outcomes varied globally. Key model input parameters are shown in Table 1. In the cohort, COVID-19 patients were characterized as: (1) severe, and (2) critical. These proportions were sourced from current literature (12-14).

**Table 1:**
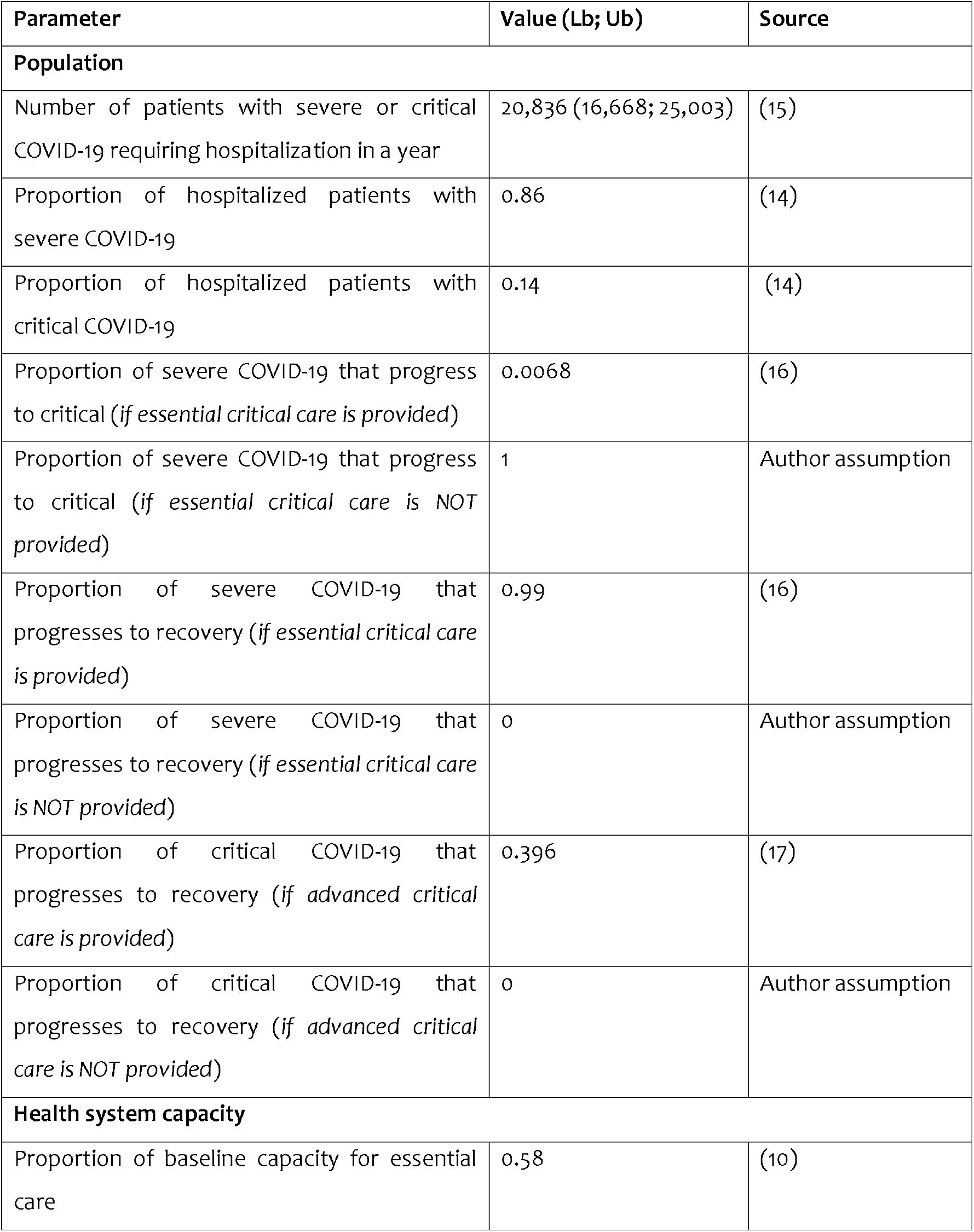
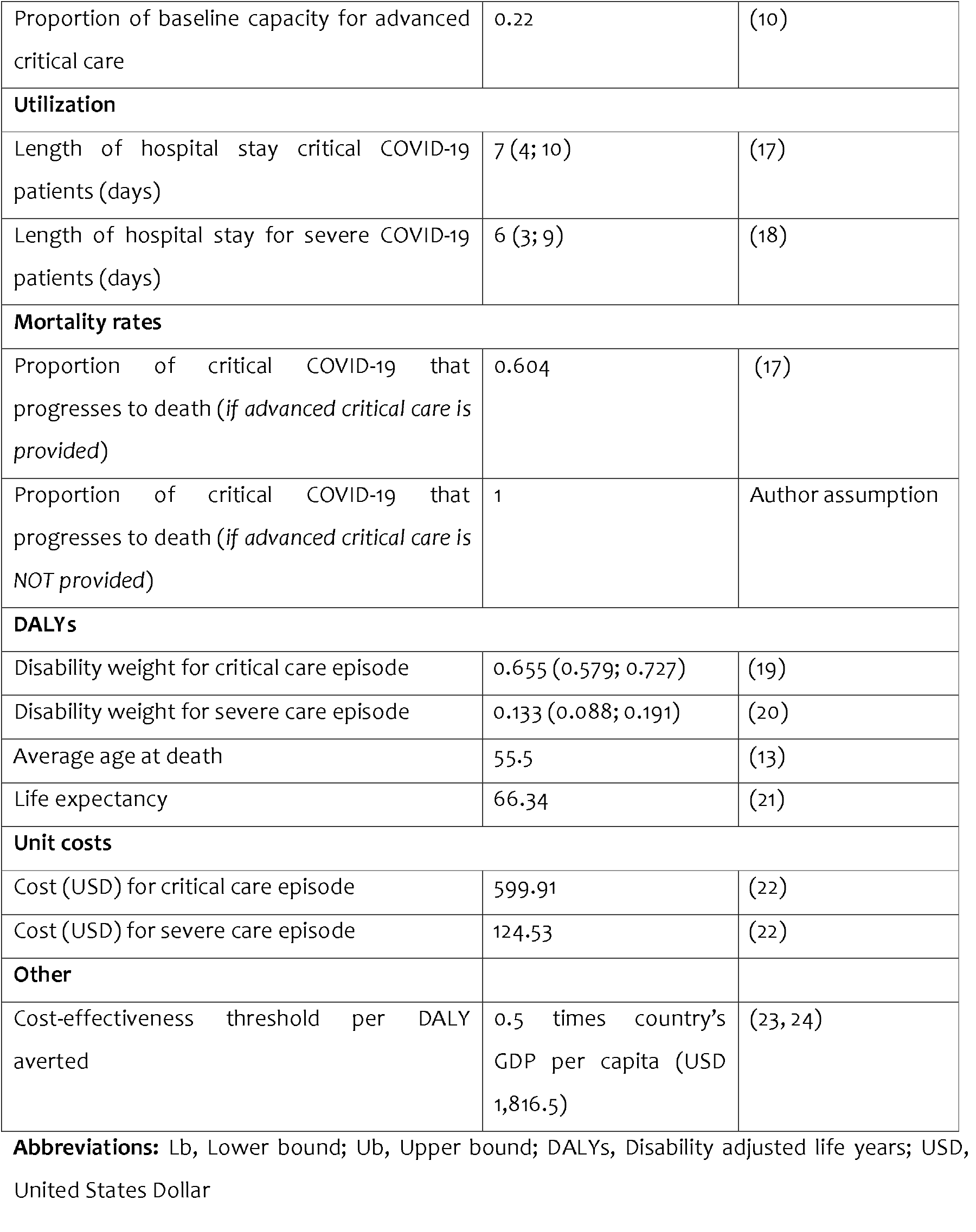
Key cost-effectiveness model parameters

### Costing methods

An ingredients-based costing methodology was used to estimate unit costs of COVID-19 case management. The health system costs considered were associated with COVID-19 case management in hospitals (accommodation and overheads, staff, pharmaceuticals, non-pharmaceutical, personal protective equipment (PPEs), oxygen therapy, ICU equipment, COVID-19 test, other laboratory tests, and radiology tests). Details of the costing and results are reported elsewhere (22).

### Effectiveness and cost-effectiveness measurement

The model’s primary outcome measure is the cost per disability adjusted life years (DALYs) averted. DALYs were calculated as the sum of years of life lost (YLL) and years of life with disability (YLD). We used standard methods to compute DALYs (25). DALYs were calculated using a discount rate of 3%, age weighting, Kenya’s life expectancy of 66.34 (21), and assumed duration of illness of 12 days. The applied disability weight for severe respiratory infection was 0.133 (0.088-0.190, 95% CI) from the global disease burden (GDB) study 2013 (20) for severe COVID-19 disease, and the disability weight of 0.655 (0.579-0.727) for ICU admission (19) for critical COVID-19 disease.

The incremental cost-effectiveness ratio (ICER) was the measure of cost-effectiveness calculated as the net change in total costs and DALYs averted between providing essential services compared to proving critical care for COVID-19 cases.

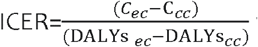 where the C_ec_ is the total cost of essential care for severe cases and C_cc_ is the total of cost of critical care

The ICER was compared to the Kenya cost-effectiveness threshold estimated by Woods and colleagues(26) (ref) and Ochalek and colleagues (27)which translated to 50% of the country’s GDP per capita (23).

### Dealing with uncertainty

A one-way sensitivity analysis was conducted across all parameters to assess the effect of changes on the ICER. A 20 % increase or decrease was implemented for the parameters. However, where possible ranges for sensitivity analysis were based on upper and lower confidence intervals or interquartile ranges found within the systematic literature review. A probabilistic sensitivity analysis (Monte Carlo simulation) was performed to explore the effect of uncertainty across our model parameters. The key parameters included the per day costs for severe and critical patients, DALYs, length of stay, and the transition probabilities with defined distributions (Appendix 2). The analysis randomly sampled each parameter in our model simultaneously from their probability distribution and repeated this 1,000 times to generate confidence intervals around our estimates of cost per DALY averted. The confidence intervals or variation of parameters and the effect on the cost effectiveness were also evaluated.

Finally, threshold analysis was run to estimate the percentage change in parameters that would render essential care and advanced critical care cost-effective using the CET as the cut-off for this determination.

## RESULTS

Table 2 shows the costs, DALYs and the incremental cost-effectiveness ratio (ICER) associated with the three analysis options. The findings show that investing to fill capacity gaps in both essential care and advanced critical care is the most costly option, followed by the status quo option. Investment to fill gaps in essential care is the least costly option. Further, investments to fill capacity gaps in both essential care and advanced critical care is the most effective option (averts the most DALYs) while status quo option is the least effective option (averts the least DALYs). The status quo option is thus dominated by investment in essential care since it is both more costly and less effective that the later. The incremental cost effectiveness ratio (ICER) of Investment in essential and advanced critical care (EC+ACC) compared to investment in essential care (EC) is US $1,378.21 per DALY averted. This is higher than the cost-effectiveness threshold for Kenya (USD 908), revealing that it is not cost-effective to prioritize the investment in advanced critical care.

**Table 2:** Cost-effectiveness results (USD 2020)

| Strategy | Total costs (USD) (95% CI) | Total DALYs (95% CI) | Cost per DALY averted (USD) | Incremental cost per DALY averted (USD) |
| --- | --- | --- | --- | --- |
| **Essential care** | 16,197,611.92<br>(15,710,057.87;<br>16,438,588.93) | 22,508.76<br>(22,232.71;<br>24,809.92) | 719.61 |  |
| **Status quo** | 17,474,037.20<br>(17,116,393.72;<br>17,728,192.34) | 77,621.36<br>(74,977.46;<br>86,558.08) | 225.11 | -23.16 |
| **Essential care and advanced critical care** | 26,156,638.28<br>(25,594,910.55;<br>26,468,628.66) | 15,282.71<br>(15,027.06;<br>16,817.24) | 1711.52 | 1,378.21 |

**Abbreviations:** USD, United States Dollar

### Sensitivity analysis

The univariate sensitivity analyses results are presented in the tornado diagram (Figure 2a) which summarizes the results for the four main parameters that had the largest effect on the ICER. These are: 1) probability of critical COVID-19 patients to progress to death (advanced critical care is provided) (lower mortality improves cost effectiveness); 2) length of stay for critical COVID-19 patients (shorter length of stay improves cost effectiveness); 3) the cost per day for advanced critical care (less costly improves cost effectiveness); 4) years of life lost (if more lost life years can be averted, essential care and advanced critical care becomes more cost effective).

**Figure 2a:**
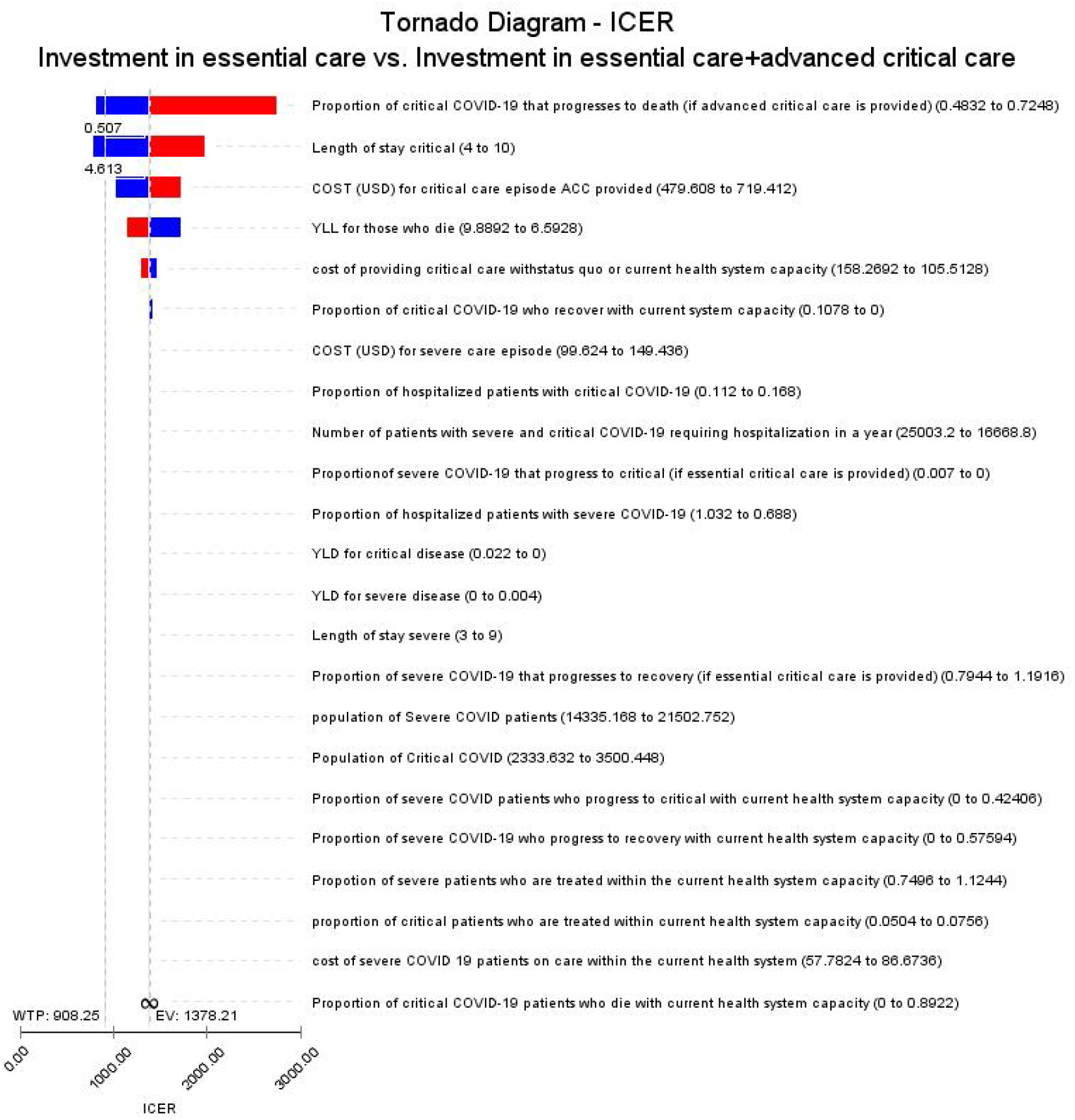
Tornado diagram of Univariate sensitivity analysis of the parameters affecting the ICER.

These results indicate that if EC + ACC strategy was less costly (by unit cost reduction or the length of stay) or more effective (through targeting patients with more years of life to lose or reduced mortality), then this strategy would be more likely to be a cost-effective use of resources.

Figure 2b summarizes the results for the main parameters that had the largest effect on the ICER comparing status quo to investment in essential care. These are: 1) length of stay for critical COVID-19 patients; 2) length of stay for severe COVID-19 patients (both of which shorter length of stay improves cost effectiveness); 3) the cost per day for essential care (less costly improves cost effectiveness); 4) probability of severe COVID-19 patients to progress to recovery (current health system capacity) (higher recovery rates improves cost effectiveness).

**Figure 2b:**
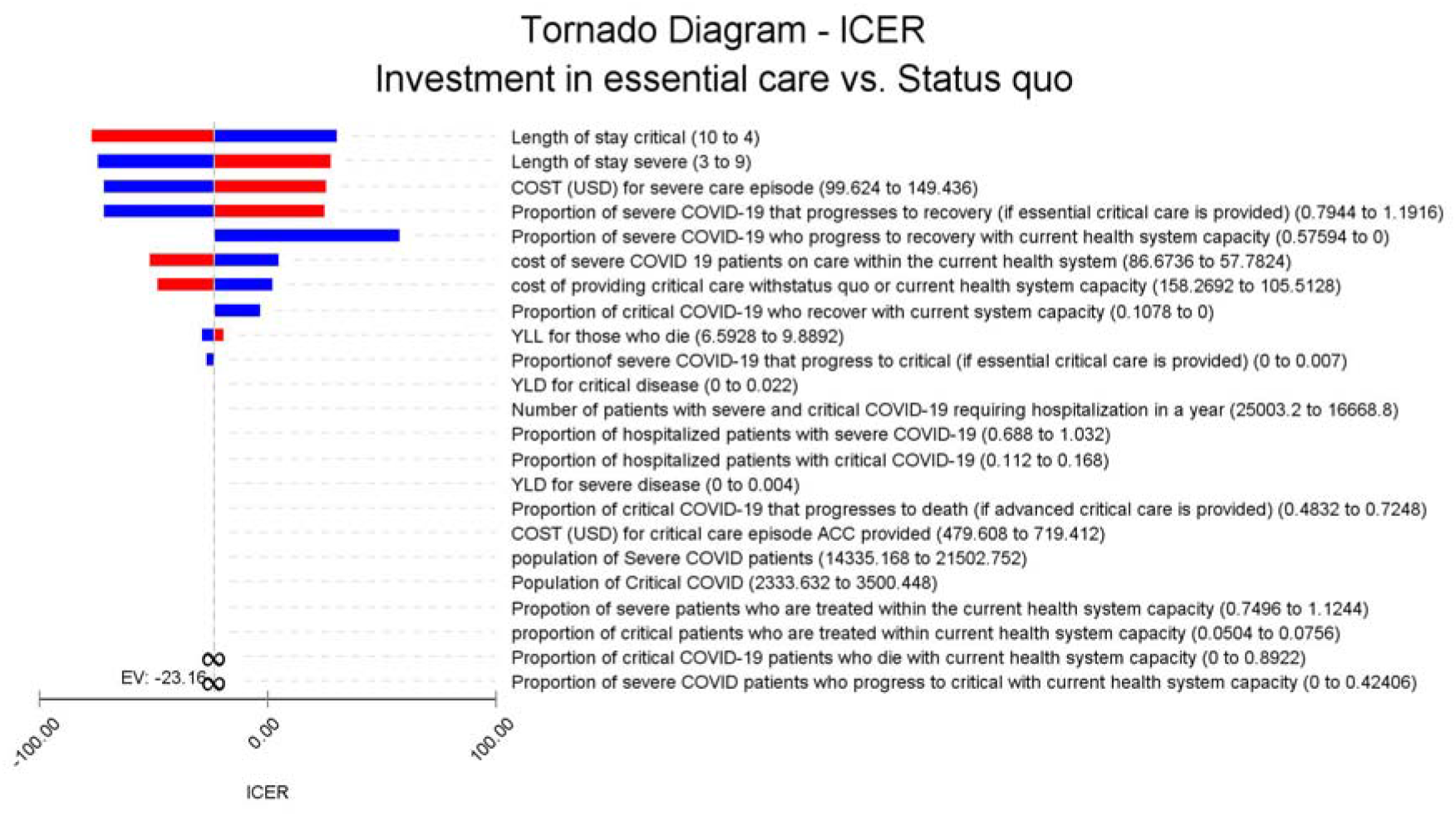
Tornado diagram of Univariate sensitivity analysis of the parameters affecting the ICER.

These results indicate that if status quo strategy was less costly (by unit cost reduction or the length of stay) or more effective (through less patients progressing to critical disease or more severe COVID 19 patients recovering), then this strategy would be more likely to be a cost-effective use of resources.

Figures 3(a-c) outline findings of the probabilistic sensitivity analysis (PSA). The region with green dots below the WTP line shows all the points that are cost effective for the EC + ACC strategy. The findings show that at a cost-effectiveness threshold of USD 908.25, the probability of EC+ACC being the more cost-effective strategy is 22%.

**Figure 3a:**
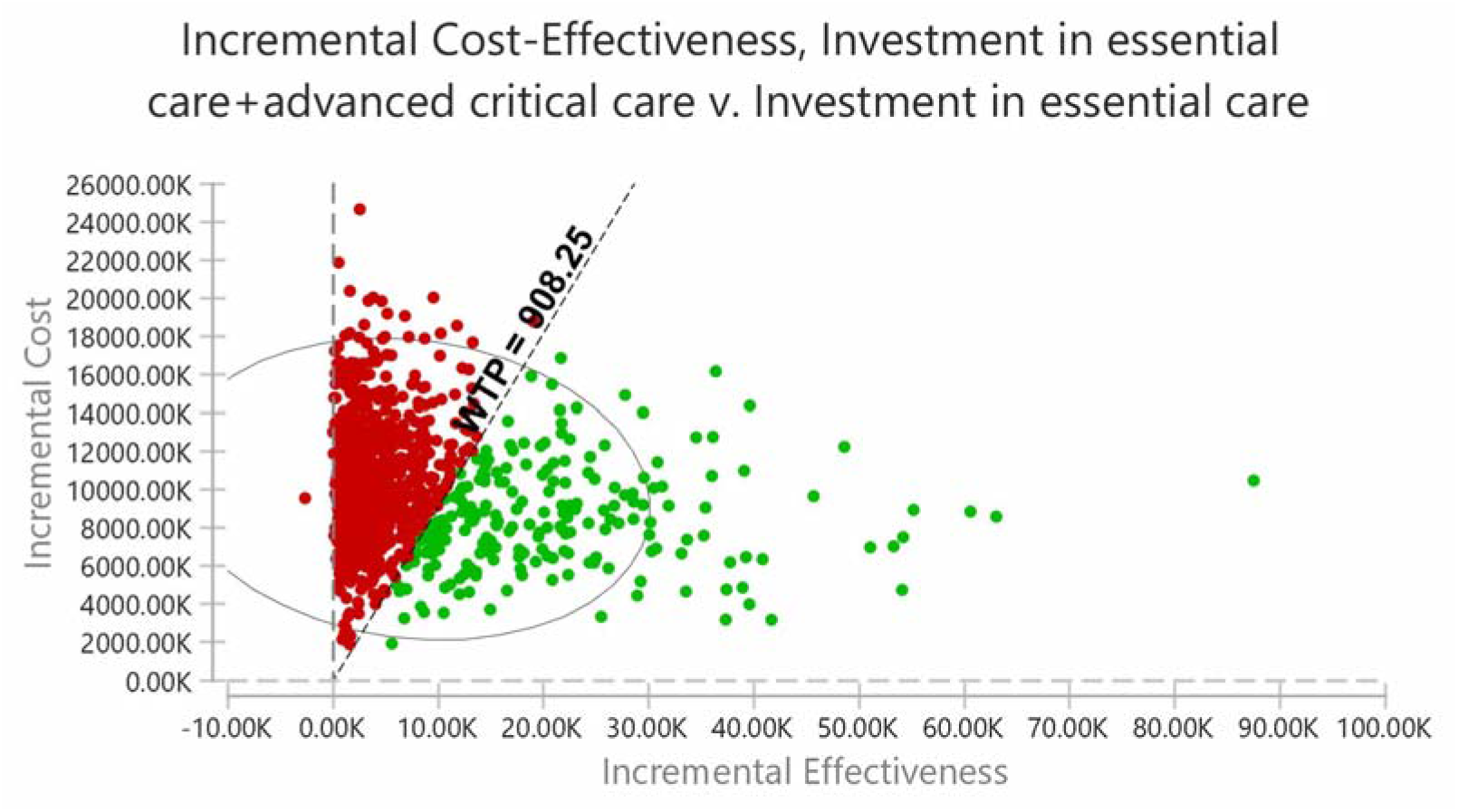
Threshold analysis of essential care versus essential and advanced critical care for COVID-19 patients A dot represents a pair of values of incremental cost and incremental effectiveness. Green dots represent the points that are cost effective (below the willingness-to-pay (WTP) threshold) Red dots represent the points that are not cost effective (above the willingness-to-pay (WTP) threshold)

**Figure 3b:**
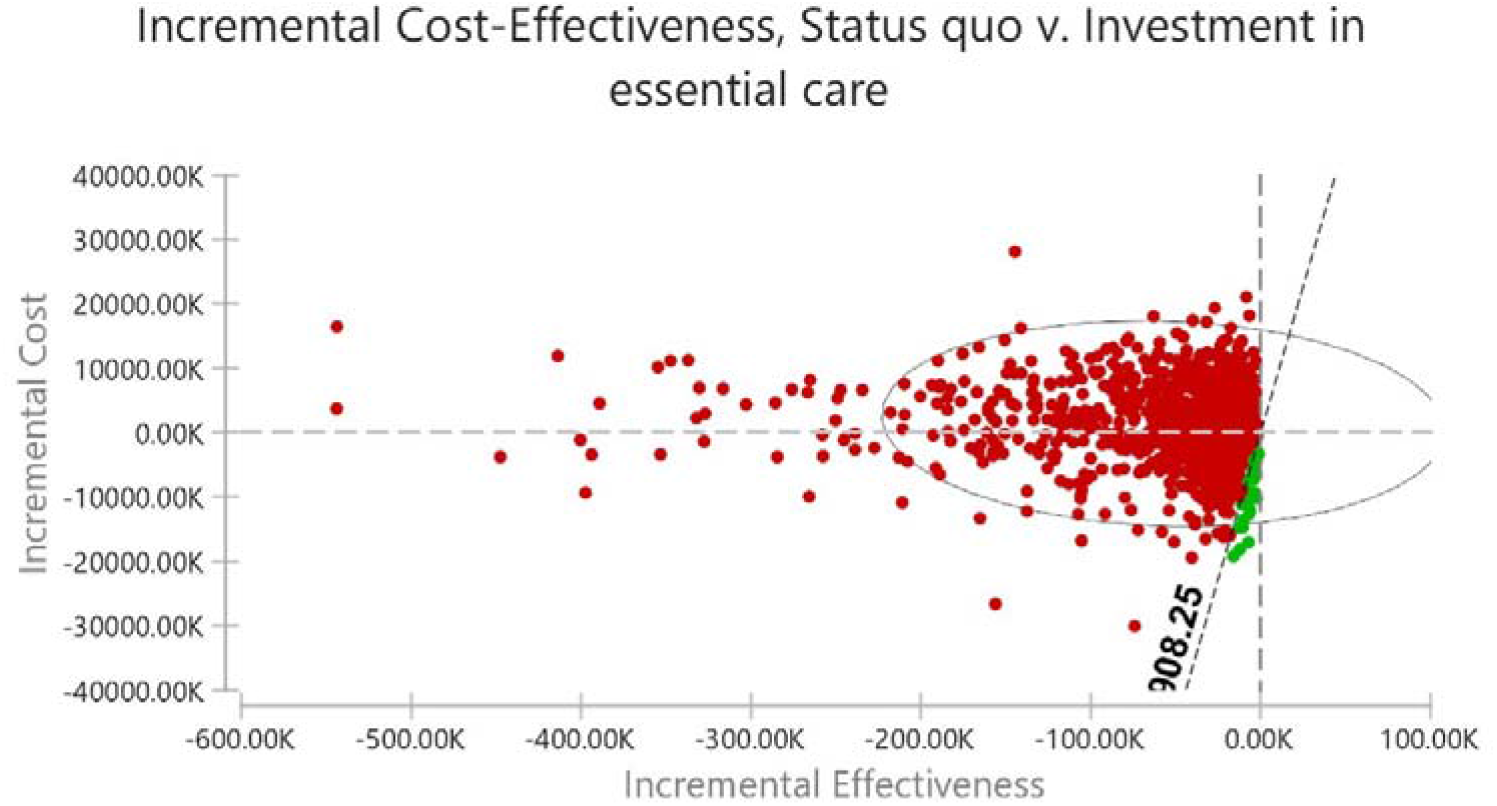
Threshold analysis of status quo versus essential care for COVID-19 patients

**Figure 3c:**
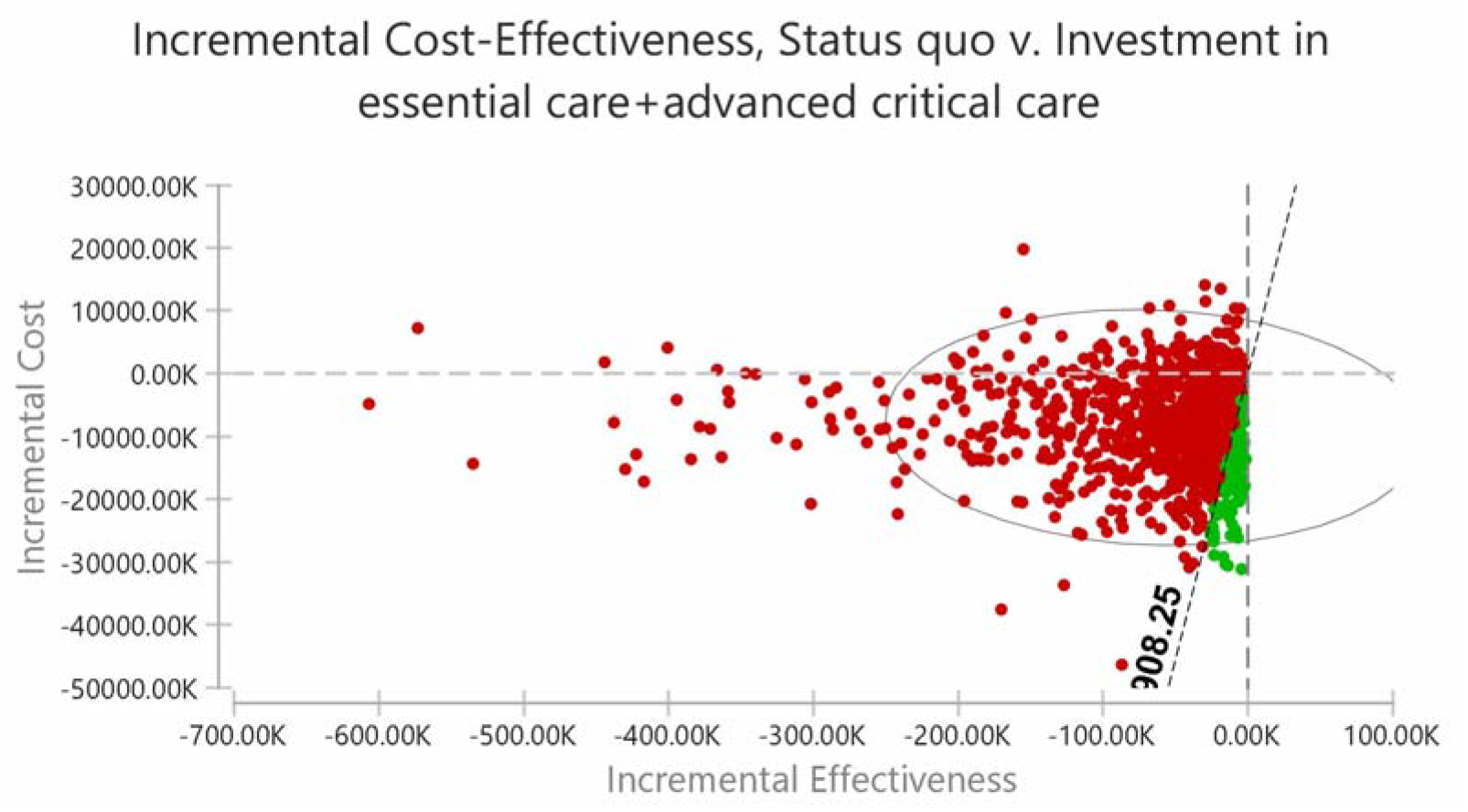
Threshold analysis of status quo versus essential and advanced critical care for COVID-19 patients A dot represents a pair of values of incremental cost and incremental effectiveness. Green dots represent the points that are cost effective (below the willingness-to-pay (WTP) threshold) Red dots represent the points that are not cost effective (above the willingness-to-pay (WTP) threshold)

The findings show that at a cost-effectiveness threshold of USD 908.25, the probability of status quo being the more cost-effective strategy is 2.7% when compared to essential care(figure 3b), and a probability of 10.8% of being the more cost-effective when compared EC + AC strategy (figure 3c).

## DISCUSSION

This study presents a cost-effectiveness analysis of investment strategies to fill gaps in Kenya’s health system capacity to provide case management for hospitalized COVID-19 patients. Specifically, we compare decisions to prioritize investments to fill existing gaps in essential care versus prioritizing to fill gaps in critical care in addition to essential care for COVID-19 patients. Our findings show that it is more cost-effective to prioritize (i.e. start with) investments to fill gaps in essential care rather than fill gaps in advanced critical care in addition to essential care. We offer several reflections on these findings.

The first, what explains this finding? The status quo option is both less effective and more costly. While it is intuitive that the status quo option is less effective, the finding that it is more costly needs some explanation. Under the status quo option, a large proportion of patients that need essential critical care miss it, and hence transition to critical care, which is more costly. Despite investment in advanced critical care in addition to essential care averting more DALYs compared to investments in essential care alone, it is substantially more expensive with cost per DALY averted being more than 2 times that of essential care. Other than the obvious high cost of advanced critical care, the proportion of hospitalized COVID-19 patients that need critical care (14%) is substantially lower than the proportion of patients that need essential care (86%) (14). Further, outcomes for advanced critical care are poor with only 39.6% recovering (17). These factors combine to make an investment in critical care in addition to essential care not cost effective. These findings mirror a similar analysis carried out in South Africa that found purchasing additional ICU care during COVID-19 surges not cost-effective (28).

Second, how should these findings be interpreted? We do not take these results to mean that Kenya should not invest in advanced critical care. Advanced critical care is evidently a vital intervention in the management of COVID-19 disease as evidenced in this analysis where it averts additional DALYs when combined with essential care, and may also have beneficial effects on the clinical management of other common conditions. However, within a context of a) substantial gaps in both essential care capacity and advanced critical care capacity and b) severe resource scarcity, what intervention should the Kenyan health sector prioritize? In other words, where should the Kenyan government start plugging the gaps? One option is to prioritize both, which within a budget constraint implies that both essential and advanced critical care will remain sub-optimal. This is indeed what we have observed in Kenya, with challenges in availability of essential care that includes oxygen and critical care persisting one year since the onset of the pandemic despite arguably a low pandemic case burden compared to for instance countries in Europe and the US (2). Our findings show that it is more cost-effective to start by prioritizing investments in plugging gaps in essential care before investing in plugging gaps in advanced care. These findings are therefore intended to inform the sequencing of investments in case management rather than the selection of either of essential or advanced critical care. Sub-optimal investment in both essential and critical care does not optimize health outcomes within a given budget.

Third, while the findings of this cost-effectiveness analysis are intended to inform priority setting for COVID-19 investments, they are to be considered within a multi-criteria decision-making framework that reflect societal values. Our study findings provide quantitative evidence to inform such as multi-criteria priority setting framework. While priority setting criteria that are based on “rule of rescue” might favour investments in both essential and advanced critical care, this is likely have high opportunity costs. A utilitarian consideration that aims to benefit the most people will favour the prioritization of essential care.

A key limitation to our analysis is the scarcity of local data to parametize the model. There is scant information on the clinical presentation, management, and outcomes of COVID-19 patients in Africa. We have however benefited from good quality data on COVID-19 surveillance in Kenya that has bridged this data gap. This notwithstanding, we have used assumptions and estimates from other settings for some of the parameters. While this may affect the validity of the findings for the Kenyan setting, the sensitivity analysis reveals that the data are largely robust to variations in these parameters. Second, this analysis considers COVID-19 as an acute condition although there is emerging evidence of long-term effects (29-31) and this has implications on the computation of DALYs. However, the information on long-terms effects is still evolving.

This study contributes to the growing body of literature on health economics analysis of COVID-19. Within the context of resource scarcity, Kenya will achieve better value for money if it prioritizes investments in essential care before investments in advanced critical care. This information on cost-effectiveness will however need to consider alongside other priority setting considerations that are informed by the values of the Kenyan society.

## Data Availability

All data relevant to the study are included in the article and are available in publicly accessible published papers. The links to the studies that contain the data are provided in the supplementary file

## Acknowledgements

We acknowledge the POETIC project team for their expertise on Essential Emergency and Critical Care (EECC) and contribution to the intervention definitions.

## Contributors

EB conceptualised the study. EB, AK, VW and SA collected data. AK, VW and EB analysed the data. AK wrote the first draft of the manuscript. All authors contributed to subsequent revisions of the manuscript.

## Disclaimer

The funders had no role in study design, data analysis, decision to publish, drafting or submission of the manuscript. The views expressed in the paper are those of the authors and not of the organisations they represent.

## Patient consent for publication

Not required.

## Ethics approval

This is a modelling study that relied on data from published and publicly available papers to parametize the model

## Patient and public involvement

Not applicable to involve patients or the public in the design, or conduct, or reporting, or dissemination plans of our research

## Provenance and peer review

Not commissioned, externally peer reviewed.

## Footnotes

Twitter: Angela Kairu @angiekairu and Edwine Barasa @edwinebarasa

## APPENDICES

## Appendix 1: Description of essential care and advanced critical care

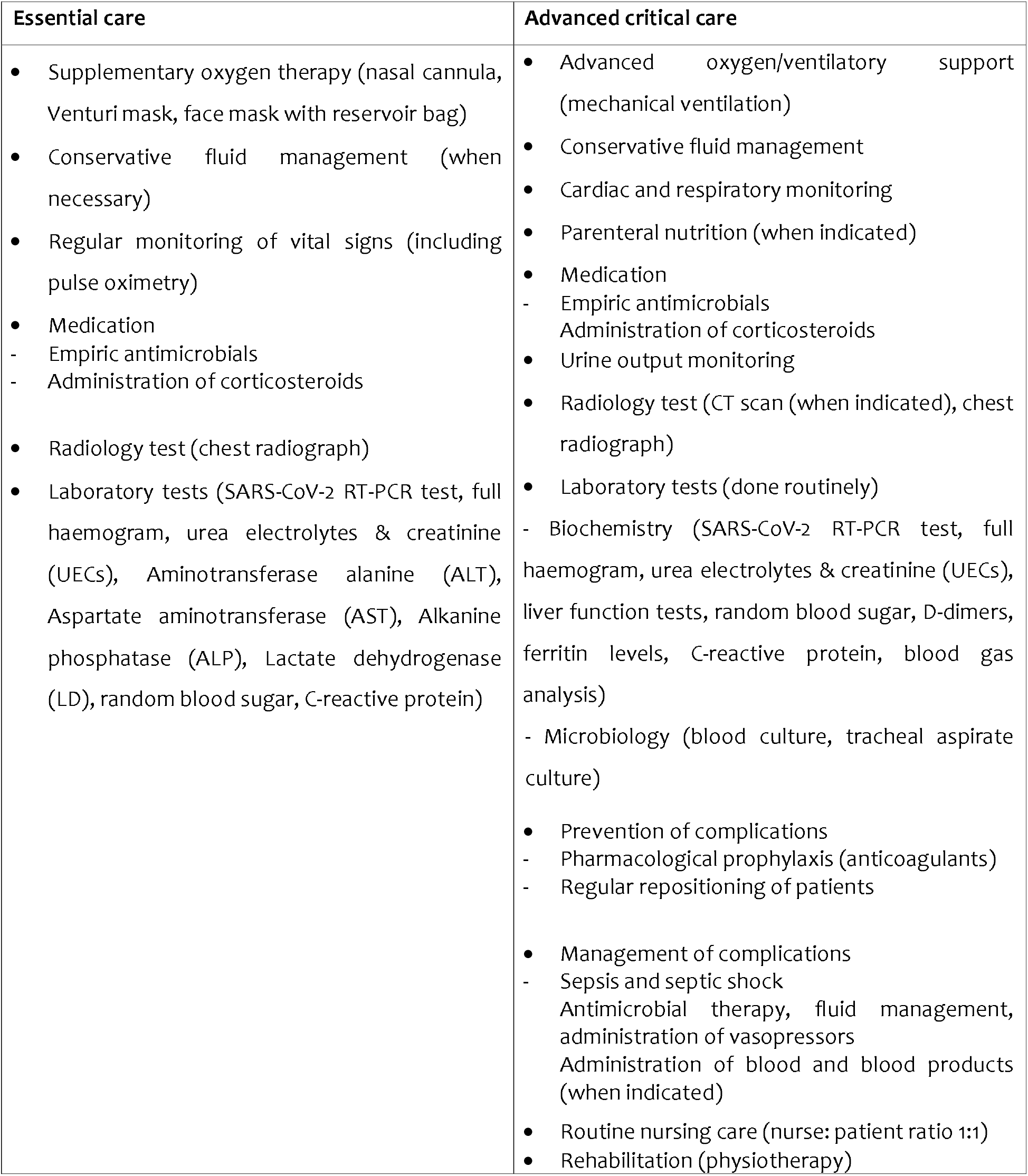

## Appendix 2: Defined distributions for model parameters

| **Model parameters** | **Distribution** |
| --- | --- |
| Total number and proportion of hospitalized patients | Normal |
| Transition probabilities | Beta |
| Costs | Gamma |
| Length of stay | Normal |
| DALYs | Lognormal |

