## Supplemental Table 1 for "Modelling the cost-effectiveness of essential and advanced critical care for COVID-19 patients in Kenya"

**Supplementary file 1**

| **Parameter** | **Value (Lb; Ub)** | **Source** |
| --- | --- | --- |
| **Population** | | |
| Number of patients with severe or critical COVID-19 requiring hospitalization in a year | 20,836 (16,668; 25,003) | [COVID-19 Transmission Dynamics Underlying Epidemic Waves in Kenya \| medRxiv](https://www.medrxiv.org/content/10.1101/2021.06.17.21259100v1) |
| Proportion of hospitalized patients with severe COVID-19 | 0.86 | [Clinical Characteristics and Outcomes of Patients Hospitalized for COVID-19 in Africa: Early Insights from the Democratic Republic of the Congo in: The American Journal of Tropical Medicine and Hygiene Volume 103 Issue 6 (2020) (ajtmh.org)](https://www.ajtmh.org/view/journals/tpmd/103/6/article-p2419.xml?rskey=XIdyGs&result=3) |
| Proportion of hospitalized patients with critical COVID-19 | 0.14 | [Clinical Characteristics and Outcomes of Patients Hospitalized for COVID-19 in Africa: Early Insights from the Democratic Republic of the Congo in: The American Journal of Tropical Medicine and Hygiene Volume 103 Issue 6 (2020) (ajtmh.org)](https://www.ajtmh.org/view/journals/tpmd/103/6/article-p2419.xml?rskey=XIdyGs&result=3) |
| Proportion of severe COVID-19 that progress to critical (*if essential critical care is provided*) | 0.0068 | [Risk factors for developing into critical COVID-19 patients in Wuhan, China: A multicenter, retrospective, cohort study - EClinicalMedicine (thelancet.com)](https://www.thelancet.com/journals/eclinm/article/PIIS2589-5370(20)30215-7/fulltext) |
| Proportion of severe COVID-19 that progress to critical (*if essential critical care is NOT provided*) | 1 | Author assumption |
| Proportion of severe COVID-19 that progresses to recovery (*if essential critical care is provided*) | 0.99 | [Risk factors for developing into critical COVID-19 patients in Wuhan, China: A multicenter, retrospective, cohort study - EClinicalMedicine (thelancet.com)](https://www.thelancet.com/journals/eclinm/article/PIIS2589-5370(20)30215-7/fulltext) |
| Proportion of severe COVID-19 that progresses to recovery (*if essential critical care is NOT provided*) | 0 | Author assumption |
| Proportion of critical COVID-19 that progresses to recovery (*if advanced critical care is provided*) | 0.396 | [Epidemiology, outcomes, and utilization of intensive care unit resources for critically ill COVID-19 patients in Libya: A prospective multi-center cohort study (plos.org)](https://journals.plos.org/plosone/article?id=10.1371/journal.pone.0251085) |
| Proportion of critical COVID-19 that progresses to recovery (*if advanced critical care is NOT provided*) | 0 | Author assumption |
| **Health system capacity** | | |
| Proportion of baseline capacity for essential care | 0.58 | [Assessing the hospital surge capacity of the Kenyan health system in the face of the COVID-19 pandemic (plos.org)](https://journals.plos.org/plosone/article?id=10.1371/journal.pone.0236308) |
| Proportion of baseline capacity for advanced critical care | 0.22 | [Assessing the hospital surge capacity of the Kenyan health system in the face of the COVID-19 pandemic (plos.org)](https://journals.plos.org/plosone/article?id=10.1371/journal.pone.0236308) |
| **Utilization** | | |
| Length of hospital stay critical COVID-19 patients (days) | 7 (4; 10) | [Epidemiology, outcomes, and utilization of intensive care unit resources for critically ill COVID-19 patients in Libya: A prospective multi-center cohort study (plos.org)](https://journals.plos.org/plosone/article?id=10.1371/journal.pone.0251085) |
| Length of hospital stay for severe COVID-19 patients (days) | 6 (3; 9) | Agweyu A. IL, Aman R., Kagucia E., Mwangangi M., Kasera K., Ng'ang'a W. Surveillance and epidemiologic evaluation of COVID-19 in Kenya (SEECK) protocol (unpublished). 2020. |
| **Mortality rates** | | |
| Proportion of critical COVID-19 that progresses to death (*if advanced critical care is provided*) | 0.604 | [Epidemiology, outcomes, and utilization of intensive care unit resources for critically ill COVID-19 patients in Libya: A prospective multi-center cohort study (plos.org)](https://journals.plos.org/plosone/article?id=10.1371/journal.pone.0251085) |
| Proportion of critical COVID-19 that progresses to death (*if advanced critical care is NOT provided*) | 1 | Author assumption |
| **DALYs** | | |
| Disability weight for critical care episode | 0.655 (0.579; 0.727) | [Assessing disability weights based on the responses of 30,660 people from four European countries \| Population Health Metrics \| Full Text (biomedcentral.com)](https://pophealthmetrics.biomedcentral.com/articles/10.1186/s12963-015-0042-4) |
| Disability weight for severe care episode | 0.133 (0.088; 0.191) | [Disability weights for the Global Burden of Disease 2013 study - The Lancet Global Health](https://www.thelancet.com/journals/langlo/article/PIIS2214-109X(15)00069-8/fulltext) |
| Average age at death | 55.5 | [EPIDEMIOLOGICAL AND CLINICAL CHARACTERISTICS OF COVID-19 PATIENTS IN KENYA \| medRxiv](https://www.medrxiv.org/content/10.1101/2020.11.09.20228106v1) |
| Life expectancy | 66.34 | <https://data.worldbank.org/indicator/SP.DYN.LE00.IN?locations=KE> |
| **Unit costs** | | |
| Cost (USD) for critical care episode | 599.91 | [Examining unit costs for COVID-19 case management in Kenya \| BMJ Global Health](https://gh.bmj.com/content/6/4/e004159) |
| Cost (USD) for severe care episode | 124.53 | [Examining unit costs for COVID-19 case management in Kenya \| BMJ Global Health](https://gh.bmj.com/content/6/4/e004159) |
| **Other** |  |  |
| Cost-effectiveness threshold per DALY averted | 0.5 times country’s GDP per capita (USD 1,816.5) | [What next after GDP-based... \| Gates Open Research](https://gatesopenresearch.org/articles/4-176/v1)  <https://data.worldbank.org/indicator/NY.GDP.PCAP.CD?locations=KE> |
